# DNA Methylation-Based Classification of CNS Tumors: Comparable Performance Between Nanopore and EPIC Technologies

**DOI:** 10.1101/2025.11.02.25339324

**Authors:** Rania Alanany, Shimaa Sherif, Apryl Sanchez, Abdulrahman Salhab, Heba Saadah, Eiman I. Ahmed, Erdener Ozer, Ata Maaz, Thidathip Wongsurawat, Noha A. Yousri, Christophe M. Raynaud, Wouter Hendrickx

## Abstract

**Background:** DNA methylation profiling enables precise classification of pediatric central nervous system (CNS) tumors. Oxford Nanopore Technologies (ONT) offers same-day, single-sample methylation readouts, but its concordance with Illumina EPIC arrays in routine diagnostic tasks remains incompletely defined.

**Methods:** We profiled 23 pediatric tumors (18 CNS, 5 non-CNS) by EPIC (v1/v2; FFPE w/o FF) and ONT (FF). CNS tumors from both platforms were classified with the crossNN_brain model; ONT data were additionally classified with Rapid-CNS2 and Sturgeon. Non-CNS tumors were classified with the crossNN pan-cancer model. We compared (i) classifier agreement with integrated histology (w/o NGS) at family/class levels, (ii) pass-rate above platform-specific score cutoffs, (iii) cross-platform concordance of copy-number variation (CNV), and *MGMT* promoter methylation status.

**Results:** In CNS cases, ONT and EPIC methylation profiles demonstrated strong correlation, except for a single low-cellularity outlier (P2), which was excluded from further analysis. Comparative assessment of the two platforms showed that: (a) Molecular classification of CNS tumors using the crossNN classifier was consistent with histology (w/o NGS) at the family level in all cases (100%, 17/17). At the class level, classification agreement was 100% (17/17) for EPIC arrays and 88% (15/17) for ONT data. (b) Copy-number profiles showed high concordance between platforms. (c) MGMT promoter methylation status matched in 94% of cases (16/17).

When comparing ONT-specific analysis pipelines using the ONT data, the Rapid-CNS2 pipeline yielded the most reliable class level assignments with 94% (16/17) concordance with the histopathological diagnosis, which marginally exceeded the crossNN classifier, while a third tool (sturgeon) underperformed. In non-CNS tumors, the pan-cancer model produced low-confidence outputs with poor agreement with histology (w/o NGS) (only 1/5 concordant), indicating limited readiness for these entities.

**Conclusions:** ONT enables same-day, clinically reliable family-level CNS tumor classification with high concordance to arrays, while EPIC retains a modest class-level edge. High concordance for *MGMT* and CNV further supports an ONT-first workflow in most CNS cases. Limitations of our study include mostly the cohort size. A key limitation of ONT is its reliance on fresh-frozen DNA and on classifiers originally built around array-derived CpG sites, rather than on models developed natively from ONT data. Building ONT-specific models could further improve class-level accuracy and confidence.

## Introduction

Pediatric central nervous system (CNS) tumors demand precise molecular diagnosis to guide therapy and prognosis, yet traditional histopathology often falls short, especially for heterogeneous or ambiguous cases [1,2]. This challenge has been recognized by the 2021 WHO CNS tumor classification, which now mandates molecular and epigenetic markers – particularly genome-wide DNA methylation profiles – for accurate diagnosis [1–3]. DNA methylation profiling, which captures the distinctive CpG methylation signatures of tumor cells, has thus emerged as a powerful complementary tool to conventional diagnostics.

Most methylation-based classification pipelines to date rely on Illumina Infinium methylation arrays (HM 450, EPIC v1 and EPIC v2). The Infinium HumanMethylationEPIC v1 BeadChip (covering ∼850K CpGs) and its updated version EPIC v2 (∼930K CpGs) provide cost-effective, high-throughput methylation measurements across gene promoters, enhancers, and other regulatory regions [4]. Notably, EPIC v2 expands enhancer coverage and supports very low-input DNA (as little as ∼1 ng) from diverse populations [4]. High-dimensional array data are then interpreted by machine-learning classifiers trained on large reference cohorts [5]. The DKFZ/Heidelberg CNS tumor classifier (random-forest based) was pioneered by Capper et al. and is now becoming routine in neuro-oncology [6–10]. Several studies have shown that array-based methylation classification has substantially improved diagnostic precision [11–14]. For example, Aldape et al. found that a high-confidence array classifier score was achieved in about two-thirds of referral cases, and that applying the classifier led to a change in diagnosis in nearly half of these high-confident cases [12]. Thus, methylation arrays and established classifiers can resolve many cases that are ambiguous by histology alone.

In parallel, Oxford Nanopore Technologies (ONT) sequencing has emerged as a promising alternative technology for methylation-based classification. ONT devices (e.g. MinION, PromethION) produce long single-molecule reads on a low-cost platform and detect 5-methylcytosine directly in native DNA without bisulfite conversion [15–18]. Crucially, ONT generates data in real time, so sequencing data (including methylation calls) can be obtained within hours rather than days [5,15–17,19]. Recent proof of concept studies illustrate this potential: Vermeulen et al. developed a neural-network (“Sturgeon”) trained on array data that could classify CNS tumors from shallow nanopore data in ∼60– 90 minutes [5,15]. In that study, models retrospectively applied to nanopore data correctly classified 45 of 50 CNS tumors using only the equivalent of 20–40 minutes of sequencing [20]. Likewise, a targeted nanopore workflow (“Rapid-CNS²”) has shown that copy-number and methylation profiles can be obtained from brain tumor specimens within about 30 min. of sequencing [21]. Importantly, these ONT-based classifiers can work with very sparse coverage (often only 1,000–10,000 CpGs) [22–24], demonstrating that even low-pass nanopore data can be sufficient for robust tumor identification.

Beyond Sturgeon and Rapid-CNS², crossNN is a newly introduced platform with explainable neural-network framework designed specifically to classify tumors from sparse, cross-platform DNA methylomes, including Illumina arrays and ONT data. crossNN was trained and validated on large multi-institutional cohorts and shown to generalize across platforms and depths, achieving high diagnostic accuracy while remaining computationally efficient and auditable (“explainable”) for clinical use. Notably, the framework supports pan-cancer prediction across >170 tumor entities spanning many organ sites, addressing classes that extend beyond the CNS space and enabling a single model to operate on both array- and ONT-derived inputs [22].

Critically, our motivation is turnaround time. Unlike array workflows that are typically run in cost-efficient batches (often ∼8 samples), and this may introduce delays while cases accumulate, the ONT workflow operates one sample at a time and returns results within clinically meaningful timeframes. To assess whether this operational advantage compromises accuracy, we performed a comparison of EPIC versus ONT on matched specimens using crossNN, which accepts both array- and ONT-derived methylomes and thus enables a fair, single-model platform comparison. In parallel, we benchmarked ONT based classifiers Sturgeon (ultra-rapid intraoperative CNS classification) and Rapid-CNS² (targeted ONT with integrated CNV/SNV/methylation readouts) to examine how algorithm design and data requirements shape diagnostic performance and time-to-answer on the same cases. Together, these analyses test whether ONT’s single-sample, rapid-turnaround workflow can be achieved without sacrificing diagnostic accuracy and clarify when each ONT-enabled approach is most appropriate.

## Material and Method

### Cohort

We conducted a retrospective, single-center study using the Sidra Pediatric Cancer Biobank. We identified 23 pediatric patients (age at diagnosis <15 years) with a final pathology diagnosis at Sidra Medicine, Qatar, between January 2018 and December 2024. Of these, 18 had central nervous system (CNS) tumors and 5 had non-CNS tumors (**Figure 1**). All patients were enrolled under protocols approved by the institutional review boards (IRB1509000) allowing the use of tumor materials for research use including diagnostic procedures and molecular analysis such as methylation profiling.

**Figure 1:**
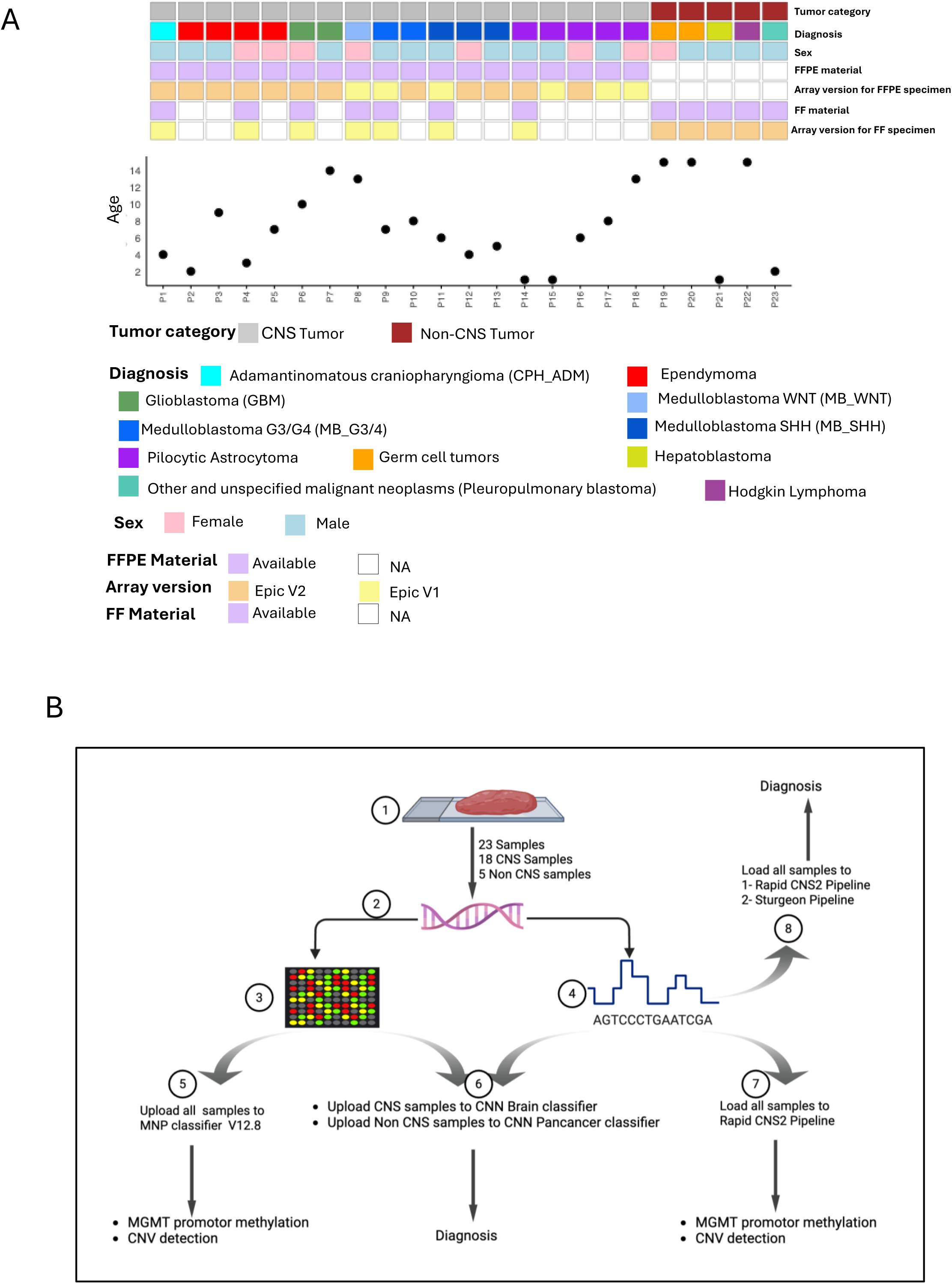
Patient cohort and analysis workflow. (A) Cohort overview of the 23 pediatric patients profiled by ONT sequencing and EPIC array, comprising 18 CNS tumors (1 adamantinomatous craniopharyngioma, 4 ependymomas, 2 glioblastoma, 6 medulloblastomas, 5 pilocytic astrocytomas) and 5 non-CNS tumors (2 germ cell tumors, 1 hepatoblastoma, 1 Hodgkin lymphoma, 1 undefined neoplasm). Color indicate sex; tiles denote availability of FFPE and/or FF tissue and the EPIC array version used per case (v1.0 or v2.0). All CNS cases had FFPE arrays (v1.0, n=6; v2.0, n=12); 7 CNS cases had matched FF profiled on EPIC v1.0; all non-CNS cases had matched FF profiled on EPIC v2.0. (B) Analysis workflow from DNA extraction to platform-specific pipelines: array data were analyzed with MNP classifier v12.8 and crossNN (brain or pan-cancer) classifiers; while the ONT data were analyzed with crossNN (brain or pan-cancer), Rapid-CNS², and Sturgeon. Mean ages for CNS and non-CNS cohorts are indicated in panel A.

The resected samples were either fresh-frozen (FF) and stored at −80° or preserved as formalin-fixed paraffin-embedded (FFPE). Five patients had both FFPE and FF samples. All FFPE tumor samples consisting of ≥70% tumor content determined by using hematoxylin and eosin (H&E) stained slides.

We also used CNS cancer retrospective cohort (GSE90496) from the published Heidelberg Molecular Neuropathology (MNP) classifier data as reference for hierarchical clustering and t-SNE integration of our samples.

### DNA Extraction

For FF samples DNA extraction was performed using Maxwell® RSC Tissue DNA Kit (#AS1610, Promega) on the Maxwell RSC instrument (#AS4500, Promega) following the manufacturers’ protocols. For FFPE samples, DNA was extracted from six 10 µm thickness curls, either immediately or after storage at -80°C for extraction on the following day, using the Maxwell® RSC DNA FFPE Kit (#AS1450, Promega).

DNA quantification was performed using the Quantus Fluorometer (#E6150, Promega) with the QuantiFluor® ONE dsDNA System (#E4870, Promega).

### Human Methylation EPIC-v1 or v2

Genome-wide DNA methylation analysis was performed using the Illumina Infinium MethylationEPIC BeadChip v1.0 or v2.0 (Illumina, San Diego, CA), which interrogates 850,000 or 930,000 CpG sites respectively at a single-nucleotide resolution. For each sample we targeted 500 ng DNA in 12 µL (proceeding with the maximum available input up to 35 µL when limited). Genomic DNA per sample was bisulfite-converted using the EZ DNA Methylation Kit (Zymo Research, Irvine, CA) according to the manufacturer’s instructions. Converted DNA was then amplified, enzymatically fragmented, and hybridized to the BeadChips following the Illumina Infinium HD Methylation protocol. Arrays underwent single-base extension, fluorescent labeling and were scanned using the Illumina iScan System. Methylated and unmethylated signal intensity data were generated as idat files. The QC of the assay control probes was performed on the Illumina GenomeStudio software.

### Nanopore sequencing

For Nanopore sequencing, 1,500 ng of genomic DNA was taken for each sample. The Ligation Sequencing Kit SQK-LSK109 (Oxford Nanopore Technologies, United Kingdom) was used to prepare libraries. Sample preparation was carried out according to the manufacturer’s protocol (Genomic DNA by Ligation, version GDE_9063_v109_revY14Aug 2019). The libraries were sequenced on a PromethION 48 (Oxford Nanopore Technologies, United Kingdom), and the loading concentration per well was 50 pM.

### Data processing for Nanopore

Pod5 files for each sample were processed using the *epi2me* human variation workflow (V.1.9.1). Modified base calling was performed with *Dorado* (V.0.3.0) dna_r10.4.1_e8.2_400bps_hac @v4.2.0 model, reads alignment to hg38 with *minimap2* (V.2.24). Bedmethyl files were generated from aligned BAM files using *modkit* (V.0.4.1) with pileup -- combine-strands --cpg then cytosine modifications were extracted. The resultant bedmethyl files were used in the downstream analysis.

### Data pre-processing for EPIC array

Raw signal intensities were extracted from IDAT files using the *minfi* Bioconductor package (V.11.52.1). Using a median log2 intensity value < 10.5 as the cutoff, all samples were included the analysis. Samples were normalized by background correction and dye-bias correction via the *funnorm* function. Beta values, obtained using the *getBeta* function, ranged from 0 (completely unmethylated) to 1 (completely methylated). For beta values derived from the EPIC_v2 dataset, the betas *CollapseToPfx* function from the sesame package (V.1.24.0) was used to collapse the beta values by averaging probes with a common probe ID prefix. During the filtration process, the following probes were removed: (a) probes targeting the X and Y chromosomes, (b) probes containing single-nucleotide polymorphisms (c) probes that cross react with more than one region in the human reference genome (hg38) [26,27], (d) probes with detection P value <0.01.

The GSE90496 reference cohort was analyzed by Illumina Infinium (H450k) and processed as previously described in Capper et.al [6].

### Comparative analysis of methylation frequencies and Visualization

Pearson correlation between methylation frequencies (from nanopore sequencing) and beta values (from the EPIC microarrays) was obtained using the *cor* function from the *stats* R package (V.4.4.1). For the unsupervised clustering, the Common probes between Sidra cohort (ONT & Arrays) and the GSE90496 cohort were extracted. Then the most variable 32,000 CpGs were filtered and the *RSpectra* (version 0.16.2) and *Rtsne* (version 0.17) were used for producing the tSNE plot [6]. However, the most variable 1,000 CpGs were filtered and the *dendextend* function from the *stats* R package (V.4.4.1) was used for the dendrogram plot.

### Heidelberg (MNP) Classifier analysis

For each sample, paired IDAT files (red and green) were uploaded into the Heidelberg DNA Methylation Tumor Classifier (version 12.8, www.molecularneuropathology.org).

The Heidelberg classifier assigned, when possible, a methylation “superfamily” and, if applicable, a more specific “family”, “class” or “subclass” within the identified superfamily, along with a calibrated score ranging from 0 to 0.99. This score indicates the degree of similarity between the sample’s methylation profile and the reference database. Scores above 0.9 are considered a match, while scores between 0.9 and 0.3 are considered as no match but indicative, as it could still be relevant for cases with low tumor content or poor DNA quality. Scores below 0.3 are considered a “no match”. Additionally, the CNV prediction and *MGMT* promoter methylation are extracted from these reports.

### crossNN classifier

For each sample, paired IDAT files (red and green) and bedmethyl file were uploaded into either the crossNN_brain classifier (for the CNS samples) or crossNN_pancancer classifier (for the non_CNS samples) for diagnosis. Diagnostic cutoffs are tumor-type and platform specific. For the crossNN brain classifier the optimal cutoffs are > 0.4 for all microarray platforms (Illumina 450K, EPIC, EPICv2) and > 0.2 for all sequencing platforms (WGBS, targeted methyl-seq, low-coverage nanopore, and whole-genome sequencing (WGS)). However, for the crossNN Pancancer classifier the optimal cutoffs are > 0.3 for all microarray platforms and > 0.15 for all sequencing platforms.

### Sturgeon

Sturgeon General Pipeline was installed and the bedmethyl file for each sample was used to infer the corresponding diagnosis. Classifier Score < 0.8: inconclusive result. If 0.8 <= score < 0.95: confident result that the class is correct. score >= 0.95: high confident result that the class is correct[5].

### Rapid-CNS^2^

Rapid-CNS² Pipeline was used as bash scripts. For each sample, bedmethyl file was utilized for diagnosis, CNV profile and *MGMT* promotor methylation prediction[21]. The classification threshold is > 0.15 [24].

## Results

### Cohort description and analysis pipeline

We profiled 23 patients by EPIC array and ONT sequencing: 18 with CNS tumors and 5 with non-CNS tumors. The CNS cohort comprised one adamantinomatous craniopharyngioma (P1), four ependymomas (P2–P5), two glioblastoma (P6-P7), six medulloblastomas (P8–P13), and five pilocytic astrocytomas (P14–P18). The non-CNS tumors included two germ cell tumors (P19–P20), one hepatoblastoma (P21), one Hodgkin lymphoma (P22), and one undefined neoplasm (potentially a Pleuropulmonary blastoma) (P23). The mean age was 6 years for CNS cases and 9 years for non-CNS cases **(Figure 1A)**. All CNS patients had FFPE tissue analyzed on Illumina Infinium MethylationEPIC arrays (v1.0, n=6; v2.0, n=12). In addition, seven CNS patients had matched FF tissue profiled on EPIC v1.0, and all non-CNS cases had matched FF tissue profiled on EPIC v2.0 (**Figure 1A**).

An overview of the analysis workflow is shown in Figure 1B: DNA extraction was followed by EPIC array profiling and ONT sequencing (see Methods). Array data were evaluated using MNP classifier v12.8 and the crossNN classifier (Brain or Pan-Cancer), while ONT data were evaluated with the crossNN classifier (Brain or Pan-Cancer), Rapid-CNS², and the Sturgeon pipeline (**Figure 1B**).

### Methylation concordance between ONT and EPIC platforms

Density plot for distribution of methylation values across CpG sites revealed a clear bimodal distribution with distinct peaks at the unmethylated (0%) and fully methylated (100%) states. This pattern is typical for DNA methylation data, reflecting CpG sites that are either largely unmethylated or heavily methylated (**Figure 2A**). Using the most variable 32,000 CpG sites across all samples [6], we observed strong concordance (0.75 - 0.95) between matched ONT and EPIC array profiles for CNS tumors in all but one case (P2; 0.25) (**Figure 2B**). By contrast, ONT–EPIC concordance was lower for non-CNS tumors (0.55 - 0.94) **(Figure 2B)**. As expected, EPIC methylation values generated from FF and FFPE material of the same patient were highly concordant (0.85 - 0.99) **(Figure 2C)**. Likewise, ONT methylation values correlated well with the corresponding EPIC methylation values derived from either FF tissues (0.90 – 0.95; **Figure 2D**) or FFPE (0.85 – 0.93; **Figure 2E**).

**Figure 2:**
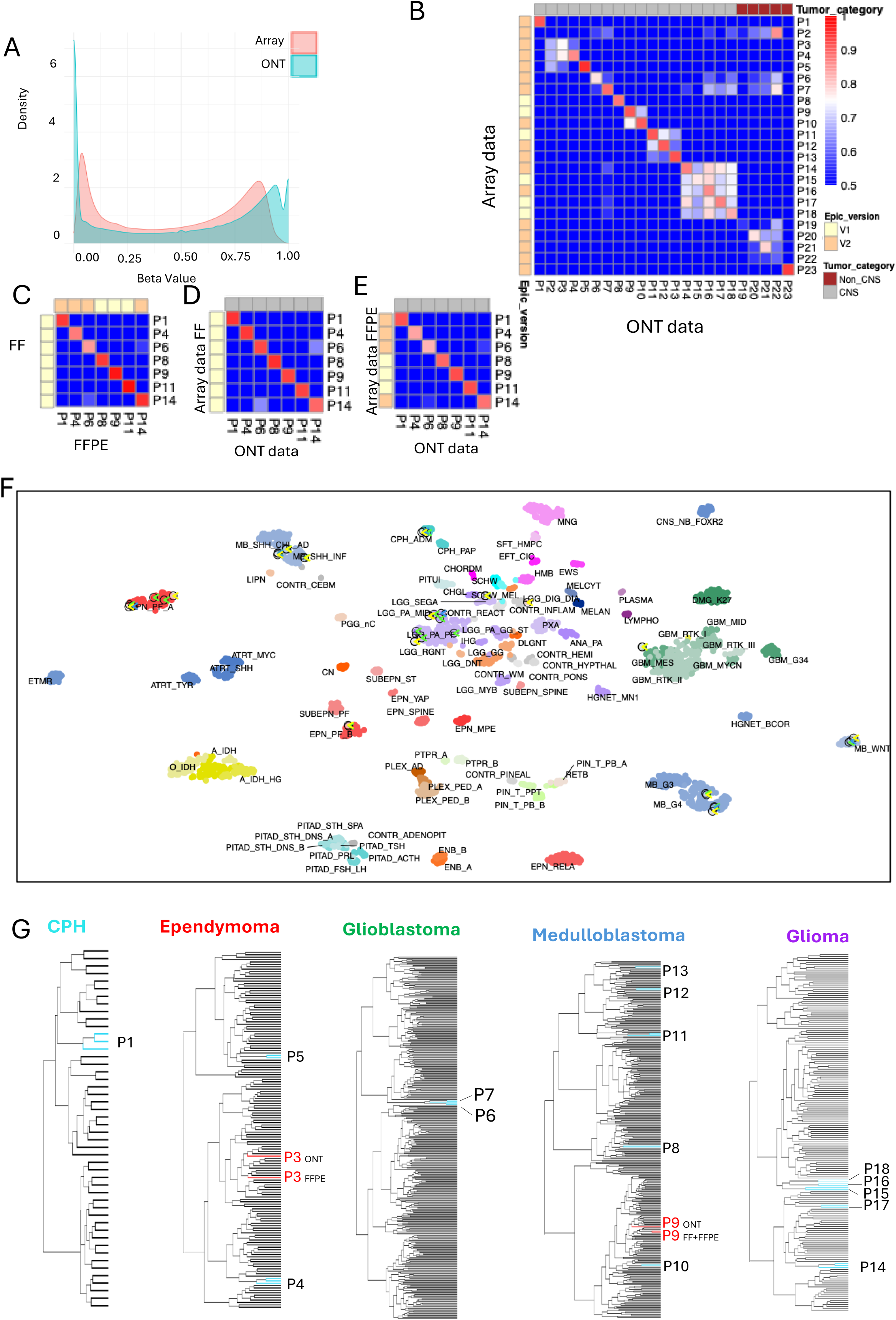
Concordance between ONT sequencing and EPIC arrays, and unsupervised clustering. (A) Distribution of methylation values: both platforms showed bimodal distribution of methylation values. (B) Correlation between matched ONT methylation frequencies and EPIC β-values using the most variable 32,000 CpGs: strong correlation for CNS tumors in all but one case (P2), with lower correlation for non-CNS tumors. (C) High within-patient correlation of EPIC β-values between FF and FFPE material. (D) ONT methylation frequencies correlate well with EPIC β-values derived from matched FF (D) and FFPE (E) tissues. (F) Unsupervised t-SNE embedding of our CNS cases together with the 2,801-sample Capper et al. reference cohort: matched specimens from each patient to the same pathology-defined subtype cluster (per-patient zooms in Supplementary Fig. 1A–R); exceptions include P2, whose ONT profile falls within the “control inflamed” cluster, and P6 (glioblastoma), which localizes near the low-grade glioma cluster. (G) Unsupervised hierarchical clustering using the most variable 1,000 CpGs within each CNS subtype (Capper reference plus our FF EPIC, FF ONT, and FFPE EPIC profiles) shows tight co-clustering of matched specimens for each patient; in two cases (P3, P9), pairs are not immediate neighbors yet remain within the same subcluster. Abbreviations: FF, fresh-frozen; FFPE, formalin-fixed paraffin-embedded; CPH, craniopharyngioma.

We next asked whether matched profiles co-localize in disease space when embedded with a large reference cohort. We therefore embedded our CNS cases together with the 2,801-sample Capper et al. reference using t-SNE, testing whether ONT and EPIC from the same patient fall into the same subtype cluster (**Figure 2F**; per-patient zooms in **Supplementary Figures 1A–R**). All EPIC and ONT pairs are localized in the same cluster. The exception was P2: the FF EPIC v1.0 profile clustered with ependymoma, whereas the matched ONT profile mapped to the “control inflamed” cluster (**Supplementary Figure 1B**). This mirrors the weak correlation results (**Figure 2B**). We hypothesize that this discordance and the reduced ONT–EPIC correlation for P2 in Fig. 2B suggest the ONT specimen likely sampled resection-margin tissue with low tumor content (likely dilution by adjacent non-neoplastic tissue), while the EPIC assays were run on a tumor-richer block. However, this remains inferential and other contributors (e.g., DNA quality, coverage depth, batch effects) cannot be excluded. Due to the discordance between ONT and EPIC methylation profiles for patient 2, this patient was excluded from further analysis.

Additionally, one pair of EPIC/ONT for patient P6 is clustered together in the same cluster (low-grade glioma cluster) which does not align with pathology-defined diagnosis of glioblastoma (**Supplementary Figure 1F)**. This discrepancy may be due to suboptimal DNA concentration in this sample (329 ng) which was lower than the average (600ng) across other samples. This was reflected in the reduced number of CpGs detected 759,524 compared to the expected ∼920,000.

As a complementary, subtype-restricted view, we performed unsupervised hierarchical clustering using the 1,000 most variable CpGs within each CNS subtype combining the Capper reference cohort [6] with our cases (**Figure 2G**). Matched specimens from the same patient clustered tightly in all but two instances P3 and P9 **(Figure 2G)**. In these cases, the paired specimens were not immediate neighbors yet remained within the same subcluster, indicating highly similar methylation profiles across the 1,000 CpGs **(Figure 2G)**.

### Comparison of molecular classification of CNS tumors using crossNN classifier

To enable a direct platform-to-platform comparison of CNS tumor classifications, we used the crossNN_brain classifier, which natively accepts both ONT and EPIC array methylation data.

Using EPIC arrays data, 16/17 samples (94%) reached a confidence score ≥0.4 at the family level, versus 13/17 (76%) at the class level (**Figures 3A and 3B**). The same analysis on ONT data is shown in **Figure 3C** (per-sample) and the results are summarized in **Figure 3D**, 15/17 samples (88%) achieved MCF score≥0.2 at the family level and 11/17 (65%) achieved MC score ≥0.2 at the class level. It is worth noting that confidence scores are platform-specific, with calibrated cutoffs of 0.2 for ONT and 0.4 for EPIC.

**Figure 3:**
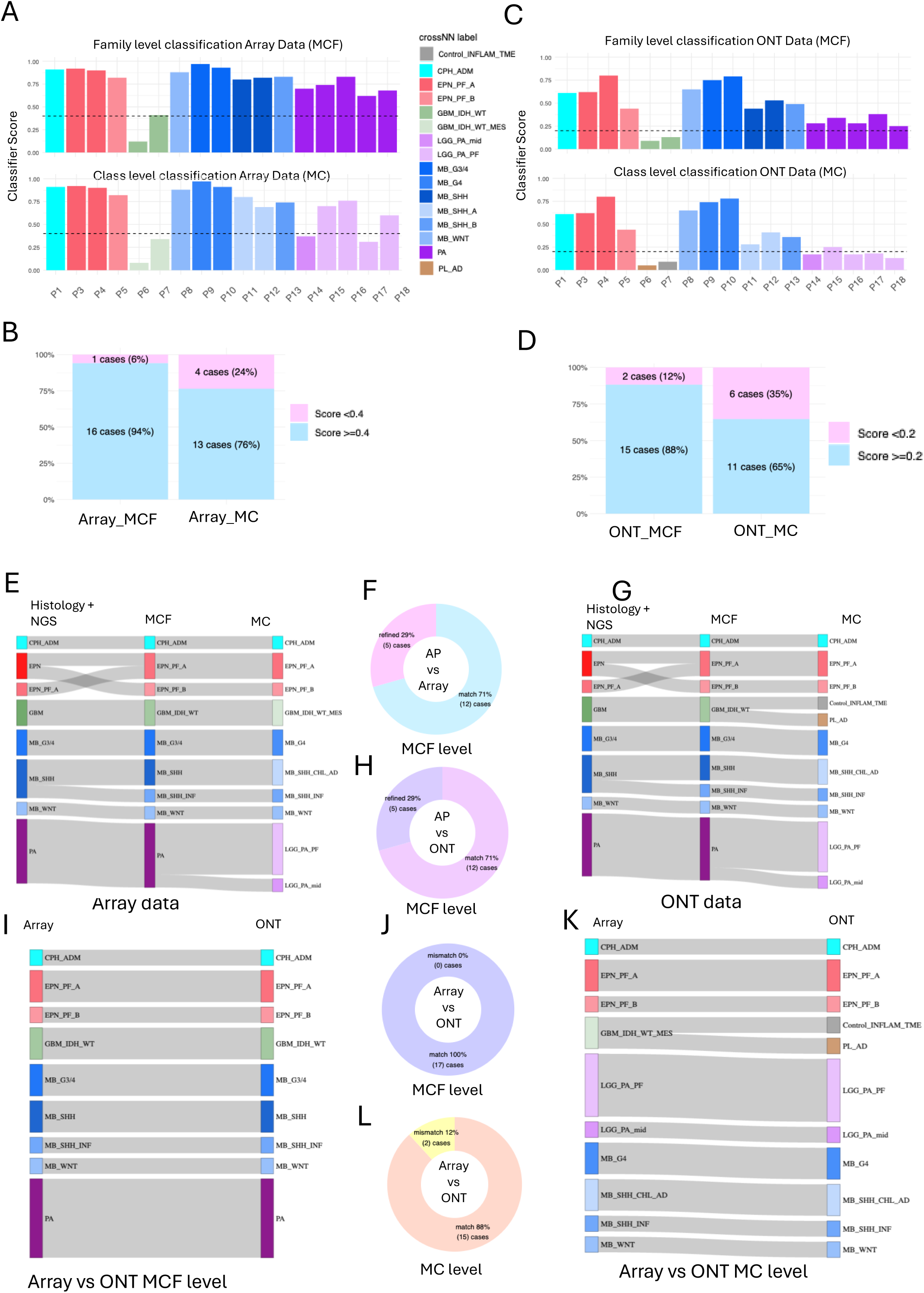
crossNN_brain classification on EPIC arrays and ONT, with integrated histology and cross-platform concordance. (A) Per-sample crossNN_brain calls from EPIC arrays, showing Methylation Class Family (MCF; top) and Methylation Class (MC; bottom) with confidence scores. (B) EPIC summary: 94% (16/17) ≥0.4 at MCF and 76% (13/17) ≥0.4 at MC. (C) Per-sample crossNN_brain calls from ONT (MCF top; MC bottom). (D) ONT summary: 15/17 (88%) ≥0.2 at MCF and 65% (11/17) ≥0.2 at MC. (E–F) Agreement of EPIC-based crossNN with histology (±NGS): 17/17 matched overall. (G–H) Agreement of ONT-based crossNN with histology (±NGS): 17/17 at MCF (H) and 88% (15/17) at MC (G). (I–J) Cross-platform concordance at the family level (MCF): ONT vs EPIC agree in 17/17. (K–L) Cross-platform concordance at the class level (MC): 15/17 agree; two tumors with low-confidence GBM family level calls diverge at the class level, being reassigned to “inflamed control” and “PL-AD,” respectively. Cutoffs are platform-specific (EPIC 0.4; ONT 0.2), and in both modalities crossNN_brain yielded a more refined molecular diagnosis in 29% cases (5/17). Abbreviations: ONT, Oxford Nanopore Technologies; EPIC, Illumina Infinium MethylationEPIC; MC, molecular class; MCF, molecular family; EPN-PF-A/B, posterior fossa ependymoma subtypes A/B; PL-AD, Plexus tumor, subclass adult.

#### Compared to integrated histology (±NGS) reference

(i) EPIC array–based crossNN calls were concordant in 17 of 17 cases (100%) at the family and class level. The crossNN classifier provided a more refined diagnosis in 5 of 17 cases (29%) at the family level (**Figures 3E and 3F**) and 12 of 17 cases at the class level (**Figure 3E)**. (ii) ONT based crossNN calls agreed with histology (±NGS) in 17 of 17 cases (100%) at the family level. Moreover, it provided a more refined molecular diagnosis in 5 of 17 (29%) (**Figures 3G and 3H**). However, at the class level (**Figure 3H**) crossNN calls refined the diagnosis in 12 of 17 cases, in addition to 2 discordant cases (**Figure 3G**).

Cross-platform concordance was high: at the family level, ONT and EPIC agreed for 17 of 17 patients (100%) (**Figures 3I and 3J**). At the class level, ONT and EPIC agreed in 15 of 17 cases (88%) (**Figures 3K and 3L**). Two samples that were concordant at the family level diverged at the class level: both tumors initially labeled as GBM at the family level (with low confidence) were reassigned to “inflamed control” and “PL-AD,” respectively.

Taken together, at the family level the ONT-based crossNN_brain classification performs essentially on par with EPIC (16/17 vs 15/17 above their respective confidence cutoffs; high cross-platform and histology concordance). By contrast, at the finer class level the EPIC array shows a modest advantage (13/17) surpassing the cutoff vs (11/17) for ONT, with higher histology agreement.

### Evaluation of molecular markers reported by both methods for the CNS tumors

crossNN does not report chromosomal copy-number profiles or *MGMT* promoter methylation status. To compare these features across platforms, we obtained copy-number calls and *MGMT* status from the Heidelberg classifier and Rapid-CNS² pipeline for EPIC array data and ONT data, respectively (**Supplementary Figure 2**). Overall, copy-number profiles were highly concordant between EPIC and ONT. Occasional differences (highlighted in blue in Supplementary Figure 2) are more plausibly attributable to multi-regional tumor sampling than to platform bias.

*MGMT* promoter methylation calls were identical between ONT and EPIC in 16 of 17 cases (94%) (**Supplementary Figure 3A**). The sole discordant case (P7) also showed low crossNN confidence on both ONT and EPIC data.

### Comparison of molecular classification of non-CNS tumors using crossNN classifier

To benchmark cross-platform performance on non-CNS tumors, we applied the pan-cancer crossNN classifier (crossNN_pancancer) to matched ONT and EPIC datasets. Using the pre-specified confidence cutoff of 0.3 for EPIC array and 0.15 for ONT data, only 1/5 samples (P23) exceeded the threshold on either platform (**Figure 4A–D**). Relative to the integrated histology diagnosis, both platforms EPIC array and ONT achieved concordance in only 1 case (P19), with confidence score 0.28 for EPIC array and 0.1 for ONT, both scores are below threshold confidence score (4/5 mismatches) **Figure 4E-H**. Cross-platform concordance was limited, with ONT and EPIC agreeing in just 2/5 cases at both family and class level (**Figure 4I-K**).

**Figure 4:**
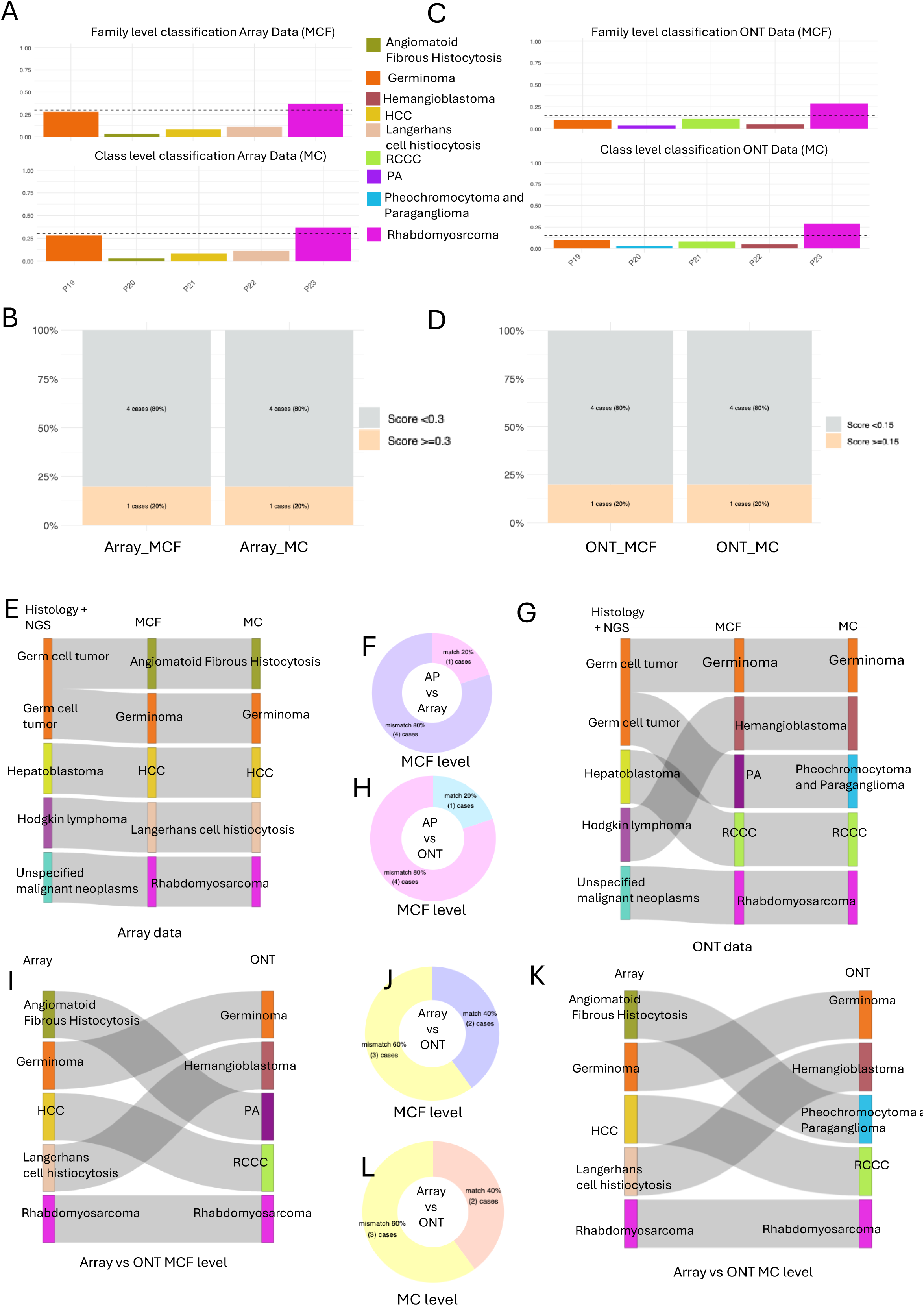
crossNN_pancancer classification on EPIC arrays and ONT, with Integrated histology and cross-platform concordance. (A) Per-sample crossNN_pancancer calls from EPIC arrays, showing Methylation Class Family (MCF; top) and Methylation Class (MC; bottom) with confidence scores. (B) EPIC summary: 20% (1/5) ≥0.3 at MCF and 20% (1/5) ≥0.3 at MC. (C) Per-sample crossNN_ pancancer calls from ONT (MCF top; MC bottom). (D) ONT summary: 20% (1/5) ≥0.2 at MCF and 20% (1/5) ≥0.2 at MC. (E–F) Agreement of EPIC-based crossNN with histology (±NGS): 1/5 matched overall; at the family levels. (G–H) Agreement of ONT-based crossNN with histology (±NGS): 1/5 matched at family (MCF) level. (I–J) Cross-platform concordance at the family level (MCF): ONT vs EPIC agree in 40% (2/5) samples. (K–L) Cross-platform concordance at the class level (MC): 40% (2/5) agreement.

To compare chromosomal copy-number profiles or *MGMT* promoter methylation status, we used as previously Heidelberg classifier for EPIC array data and Rapid-CNS² for ONT data. As for the CNS tumors we observed a good concordance with occasional differences (highlighted in blue in Supplementary Figure 2) are more plausibly attributable to multi-regional tumor sampling than to platform bias.

*MGMT* promoter methylation calls were identical between ONT and EPIC in all cases (100%), 4 unmethylated and 1 methylated (**Supplementary Figure 3B**).

In this small non-CNS cohort, the crossNN_pancancer model performed poorly, suggesting that it may be insufficiently trained/under-represented for these specific tumor entities (and/or not well calibrated for this setting). Until expanded training and calibration are available, pan-cancer outputs here should be treated as exploratory, with diagnostic decisions anchored to histology (±NGS) or organ-specific classifiers.

### Comparison of molecular classification of CNS tumors using crossNN, Sturgeon and Rapid-CNS² classifiers on ONT data

We compared the three ONT-compatible classifiers crossNN_brain, Rapid-CNS², and Sturgeon Across 17 evaluable CNS cases, Rapid-CNS² produced correct class level diagnoses with high internal confidence in 15/17. crossNN_brain achieved perfect family level agreement (17/17) and correct class level labels in 15/17, though several of those class level calls had low confidence giving a slight edge to Rapid-CNS². Sturgeon performed worst: it misclassified one case, yielded one class level assignment discordant with both Rapid-CNS² and crossNN_brain, and returned six calls with suboptimal classification as “glioma, IDH astrocytoma (A-IDH),” three with confidence <0.8 (**Figure 5**).

**Figure 5:**
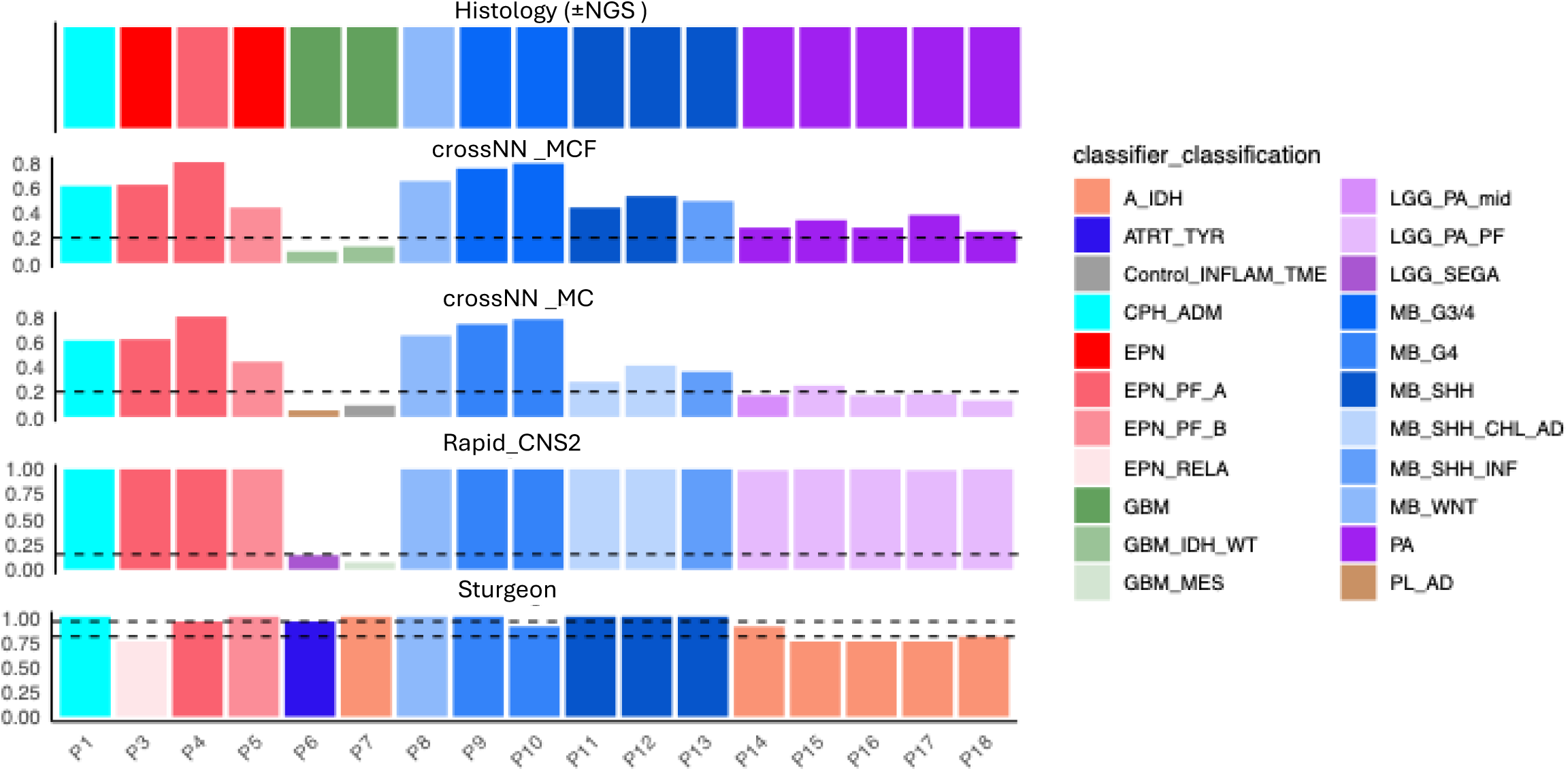
Head-to-head comparison of ONT methylation classifiers versus integrated Histology. Per-patient classifications for the 17 CNS cases profiled by ONT are shown across four rows: integrated Histology (±NGS) reference (top), crossNN_brain Methylation Class Family (crossNN_MCF), crossNN_brain Molecular Class (crossNN_MC), Rapid-CNS², and Sturgeon. Bar color encodes the assigned class; bar height encodes each tool’s internal confidence/probability. Dashed lines mark the ONT-specific crossNN cut-off (0.2) for MCF/MC; Rapid-CNS² has no formal threshold. Across the 17 evaluable cases, Rapid-CNS² produced correct class level calls with high confidence in 88% (15/17). crossNN_brain achieved perfect agreement at the family level 100% (17/17) and correct class level labels in 88% (15/17), though several class level scores fell below the 0.2 cut-off.

Overall, Rapid-CNS² marginally outperformed crossNN_brain, with Sturgeon trailing; these comparisons should be interpreted cautiously given the small cohort size.

## Discussion

Accurate, timely molecular classification is central to pediatric CNS tumor management. Using matched specimens, we benchmarked ONT sequencing against Illumina EPIC arrays with a unified analytic framework (crossNN). Methodologically, ONT provides a genome-wide readout that samples methylation at millions of CpG sites far beyond the fixed target set of arrays. The trade-off is per-site depth: in typical diagnostic runs, individual CpGs are often supported by only a handful of nanopore reads. By contrast, EPIC arrays interrogate a predefined panel of ∼0.85–0.93 million CpGs (v1–v2) with highly standardized, replicated bead-level measurements and internal controls, yielding high per-site precision. Despite these different sampling regimes, our matched profiles correlated strongly across the 32k most variable CpGs, and unsupervised tSNE consistently co-localized each patient’s ONT and EPIC profiles within the same CNS reference neighborhoods. The single clear exception (P2) behaved consistently across analyses: the ONT profile clustered with “inflamed control,” while the EPIC profile clustered with the Ependymoma subtype. This is maybe due to low tumor cellularity in the ONT specimen rather than a platform bias.

When both modalities were analyzed with crossNN_brain using platform-specific thresholds, EPIC and ONT agreed at the family level in 94% cases and at the class level in 83%. Relative to integrated pathology, EPIC matched in 100% overall, while ONT matched in 100% at family level and 88% at class level, and both modalities provided more refined labels in a subset of cases. Two practical biomarkers showed high cross-platform concordance: genome-wide copy-number profiles were largely superimposable, and MGMT promoter calls were identical in 100% (5/5) non-CNS and 94% (16/17) CNS cases, with a single discordance aligned with low classifier confidence. Together, these findings argue that ONT preserves the clinically salient methylome signal needed for classification and ancillary readouts.

Our ONT-tool comparison highlights algorithmic trade-offs. Rapid-CNS² produced the highest proportion of correct, high-confidence class level labels in our 17 evaluable CNS cases, whereas crossNN_brain achieved perfect family level agreement and the same number of correct class labels but with several below its own ONT cutoff. Sturgeon underperformed, with two misclassifications, one of them is discordant at class level. However, there is tendency toward 6 suboptimal classifications as A-IDH glioma calls.

Since finalizing our analysis, a unified ONT workflow ROBIN (Rapid nanpOre Brain IntraoperatIve classificatioN) has been reported in Neuro-Oncology [28]. ROBIN streams the same ONT run through multiple methylation-based classifiers; Sturgeon, crossNN, and random forest (RF) approach from RapidCNS2. These classifiers are integrated with CNV changes, coverage over targets and SNVs, and *MGMT* promoter methylation and candidate fusions in one graphical user interface. This enables confident diagnosis by a clinician in real time during sequencing. In practice, this ensemble design harmonizes score scales across tools, surfaces conflicts with orthogonal evidence, and drives borderline cases toward a stable consensus meaning the modest Rapid-CNS² vs crossNN discrepancies we note here are expected to diminish under an ensemble report that calibrates scores across tools and reconciles disagreements; in practice this typically yields a single consensus label or a conflict flag, so small Rapid-CNS² vs crossNN differences would not alter the final class reported or the pass/fail confidence call.

By contrast, the crossNN_pan-cancer model performed poorly across our five non-CNS pediatric tumors on both platforms, rarely exceeding confidence thresholds and showing limited agreement with histology (±NGS). Given the model’s training focus and our small, heterogeneous pediatric set, this likely reflects under-representation and calibration gaps rather than a platform effect. Until pediatric non-CNS coverage expands in training cohorts, pan-cancer outputs should be treated as exploratory adjuncts rather than diagnostic drivers in this setting.

Most current CNS methylation classifiers including the Heidelberg/MNP lineage and newer ONT-compatible tools are anchored to array-trained reference atlases (Illumina 450K/EPIC). Although implementations differ (e.g., the original MNP random forest uses ∼10k selected CpGs, whereas others retain larger array-derived subsets or simulate sparse sequencing inputs), the effective feature space remains largely defined by CpGs present on legacy arrays. EPIC v2 introduces additional probes, but many production pipelines still operate on the 450K/EPIC-shared backbone for backward compatibility; only recently have extended models begun to accommodate EPIC v2 and sequencing inputs. By contrast, ONT assays methylation genome wide. Retraining classifiers to learn features directly from ONT methylomes leveraging genome-wide CpGs and regional/phase-aware methylation patterns should reveal more informative markers and, in our cohort, may narrow the modest class-level advantage we still observe with arrays. Turnaround time remains the strongest operational argument for ONT. Array workflows are reliable but batched, multistep, and often multi-day. ONT delivers single-sample, same-day methylation with the added benefit of concurrent CNV/SV and selected SNV information from the same library. In multidisciplinary pediatric care, earlier molecular anchoring even at the family level can accelerate decisions without exhausting tissue.

Limitations include cohort size and composition (pediatric CNS-enriched, few diagnostically extreme edge cases), and the fact that ONT coverage is variable across loci and samples, placing a premium on stringent QC and platform-aware confidence thresholds. Importantly, in our implementation ONT required high-molecular-weight DNA from fresh-frozen tissue; routine FFPE compatibility remains limited today, whereas EPIC is validated and widely adopted on FFPE. Finally, broader, multi-institutional validation under CLIA/CAP-compliant SOPs will be essential to harden thresholds, quantify sensitivity/specificity across entities and ages, and standardize reporting.

In sum, ONT methylation profiling is a robust, rapid alternative to arrays for pediatric CNS tumor classification. Family-level diagnoses are essentially comparable across platforms; arrays retain a modest edge at class level, but ONT results are frequently confident and actionable, with the added benefits of speed and multi-omic breadth. Third, classifier design matters: ONT-native pipelines differ in confidence behavior and error modes, with Rapid-CNS² slightly better performing than crossNN_brain in our small set and Sturgeon trailing. Importantly, when an ONT run meets predefined quality metrics (e.g., adequate coverage/CpG call rate, coherent CNV profile) and yields high classifier confidence, the ONT label stands on its own array (EPIC) confirmation is not required to trust the result. With expanded pediatric training data, ONT-native feature sets, and continued calibration, ONT can sit alongside and in selected scenarios ahead of arrays in precision neuro-oncology.

## List of abbreviations

A-IDH: IDH glioma, subclass astrocytoma
ATRT_TYR: Atypical teratoid/rhabdoid tumor, subclass
TYR CAP: College of American Pathologists
CLIA: Clinical Laboratory Improvement Amendments
CNS: Central nervous system
CNV: Copy-number variation
CpG: 5′-cytosine-phosphate-guanine dinucleotide
crossNN: Neural-Network classifier (with crossNN_brain and crossNN_pancancer variants)
CPH_ADM: Craniopharyngioma adamantinomatous tumor
DKFZ: Deutsches Krebsforschungszentrum (German Cancer Research Center)
EPIC: Illumina Infinium MethylationEPIC BeadChip array (v1.0/v2.0)
EPN: Ependymoma
EPN-PF-A/B: Ependymoma, posterior fossa group
A/B FF: Fresh frozen
FFPE: Formalin-fixed paraffin-embedded
GBM: Glioblastoma
GBM_IDH_WT: Glioblastoma, IDH wildtype
GBM_IDH_WT_MES: Glioblastoma, IDH wildtype, subclass mesenchymal
GRCh38 / hg38: Human genome reference build 38
H&E: Hematoxylin and eosin
HCC: Hepatocellular Carcinoma
IDAT: Illumina array intensity file format
IDH: Isocitrate dehydrogenase
IRB: Institutional Review Board
LGG: Low-grade glioma
LGG_PA_mid: Low grade glioma, subclass midline pilocytic astrocytoma
LGG_PA_PF: Low grade glioma, subclass posterior fossa pilocytic astrocytoma
LGG_SEGA: Low grade glioma, subependymal giant cell astrocytoma
MB: Medulloblastoma
MB_WNT: Medulloblastoma WNT
MB_SHH: Medulloblastoma SHH
MC: Methylation Class (crossNN output)
MCF: Methylation Class Family (crossNN output)
MGMT: O^6-methylguanine-DNA methyltransferase
MNP: Molecular Neuropathology (Heidelberg) classifier web portal
NGS: Next-generation sequencing
ONT: Oxford Nanopore Technologies
PA: Pilocytic Astrocytoma
PF: Posterior fossa
PL-AD: Plexus tumor, subclass adult
QC: Quality control
Rapid-CNS²: Rapid comprehensive adaptive nanopore sequencing pipeline for CNS tumors
RCCC: Renal clear cell carcinoma
SNP: Single-nucleotide polymorphism
SNV: Single-nucleotide variant
SOP: Standard operating procedure
Sturgeon: ONT-trained deep-learning classifier (tool name)
SV: Structural variant
t-SNE: t-Distributed Stochastic Neighbor Embedding
TME: Tumor microenvironment
WGBS: Whole-genome bisulfite sequencing
WGS: Whole-genome sequencing

## Declarations

### Ethics approval and consent to participate

All human samples used in this study were collected under approval from the Sidra Medicine Institutional Review Board (IRB Protocol #150900).

### Consent for publication

“Not applicable”

### Data availability

All data produced in the present study are available upon reasonable request to the authors

## Acknowledgements

The authors would like to acknowledge the Sidra Medicine research administration team and the Omics Core Facility at Sidra Medicine for providing DNA methylation service using the Illumina Infinium MethylationEPIC BeadChip and ONT sequencing without whom we could not have performed this work.

## Author’s contribution

C.R., S.S. and E.O. selected all the samples. R.A. and C.R., performed the experiments, R.A. and C.R. performed the analysis. R.A. and C.R., drafted the manuscript and the figures. W.H., S.S., R.A., E.A, E.O. and C.R. edited the text. Conceptualization by W.H., R.A. C.R., and E.O., W.H. project supervision and coordination. All authors reviewed the manuscript.

---

*Supplementary figure 1: Per-patient t-SNE zooms show co-localization of ONT and EPIC profiles within reference CNS methylation classes*

Per-patient t-SNE zooms show co-localization of ONT and EPIC profiles (FFPE and FF when available) with Capper et al. reference cohort (n=2,801, 91 CNS methylation classes) (A–R). In nearly all cases, the three profiles from the same patient co-localize within the same pathology-defined subtype cluster. The main exception is P2, whose ONT profile falls within the “control inflamed” region while the EPIC profiles map to ependymoma consistent with low tumor cellularity in the ONT specimen. P6 appears near the low-grade glioma cluster despite pathology assignment *to glioblastoma*. Panel-to-patient mapping: A=P1, B=P2, C=P3, D=P4, E=P5, F=P6, G=P7, H=P8, I=P9, J=P10, K=P11, L=P12, M=P13, N=P14, O=P15, P=P16, Q=P17, R=P18. Axes represent t-SNE dimensions. Abbreviations: FF, fresh-frozen; FFPE, formalin-fixed paraffin-embedded; ONT, Oxford Nanopore Technologies; EPIC, Illumina Infinium MethylationEPIC.

Legend: ONT= yellow, FFPE= Green, FF= Blue

*Supplementary figure 2: Genome-wide copy-number profiles from ONT and EPIC across CNS cases (P1–P18)*

For each CNS patient (P1–P18), paired CNV plots are shown with ONT (Rapid-CNS²) on the right and EPIC array (Heidelberg/MNP classifier) on the left. Light blue ellipses highlight chromosomal aberrations detected on a single platform (platform-unique), while red ellipses mark aberrations shared by both platforms. Overall, profiles are largely concordant, with a limited number of platform-specific events.

*Supplementary figure 3: Genome-wide copy-number profiles from ONT and EPIC across non-CNS cases (P19–P23)*

For each non-CNS patient (P19–P23), paired CNV plots are shown with ONT (Rapid-CNS²) on the right and EPIC array (Heidelberg/MNP classifier) on the left. Blue circles highlight platform-unique chromosomal aberrations; red circles mark aberrations shared by both platforms. Overall, profiles are largely concordant, with a few platform-specific events.

*Supplementary figure 4: Concordance of MGMT promoter methylation between ONT and EPIC*

(A) CNS tumors: 16/17 (94%) concordant MGMT promoter methylation calls; 1 discordant. (B) Non-CNS tumors: 5/5 (100%) concordant.

