## Supplementary figures for "DNA Methylation-Based Classification of CNS Tumors: Comparable Performance Between Nanopore and EPIC Technologies"

Supplementary Figure 1

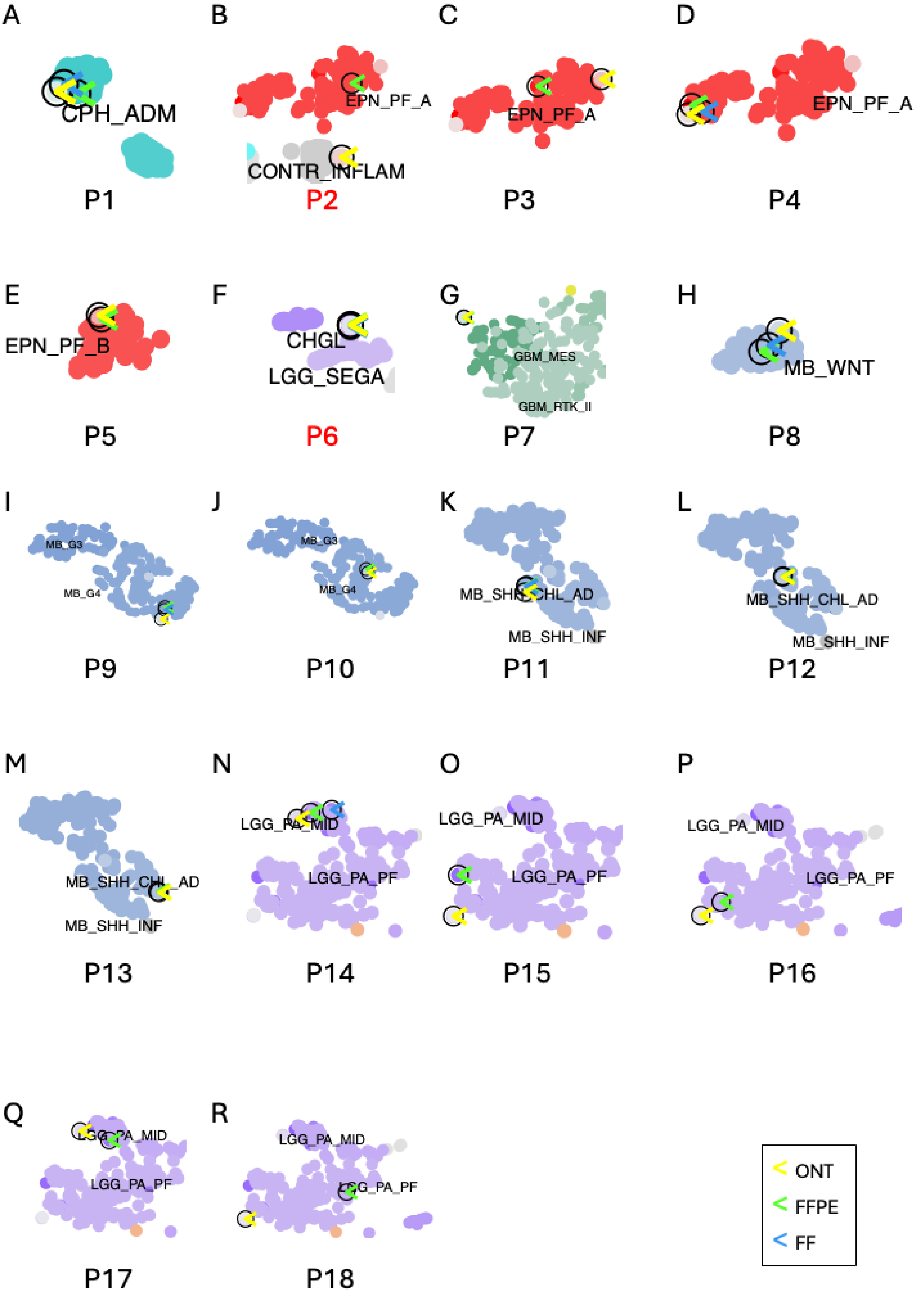

Supplementary Figure 2

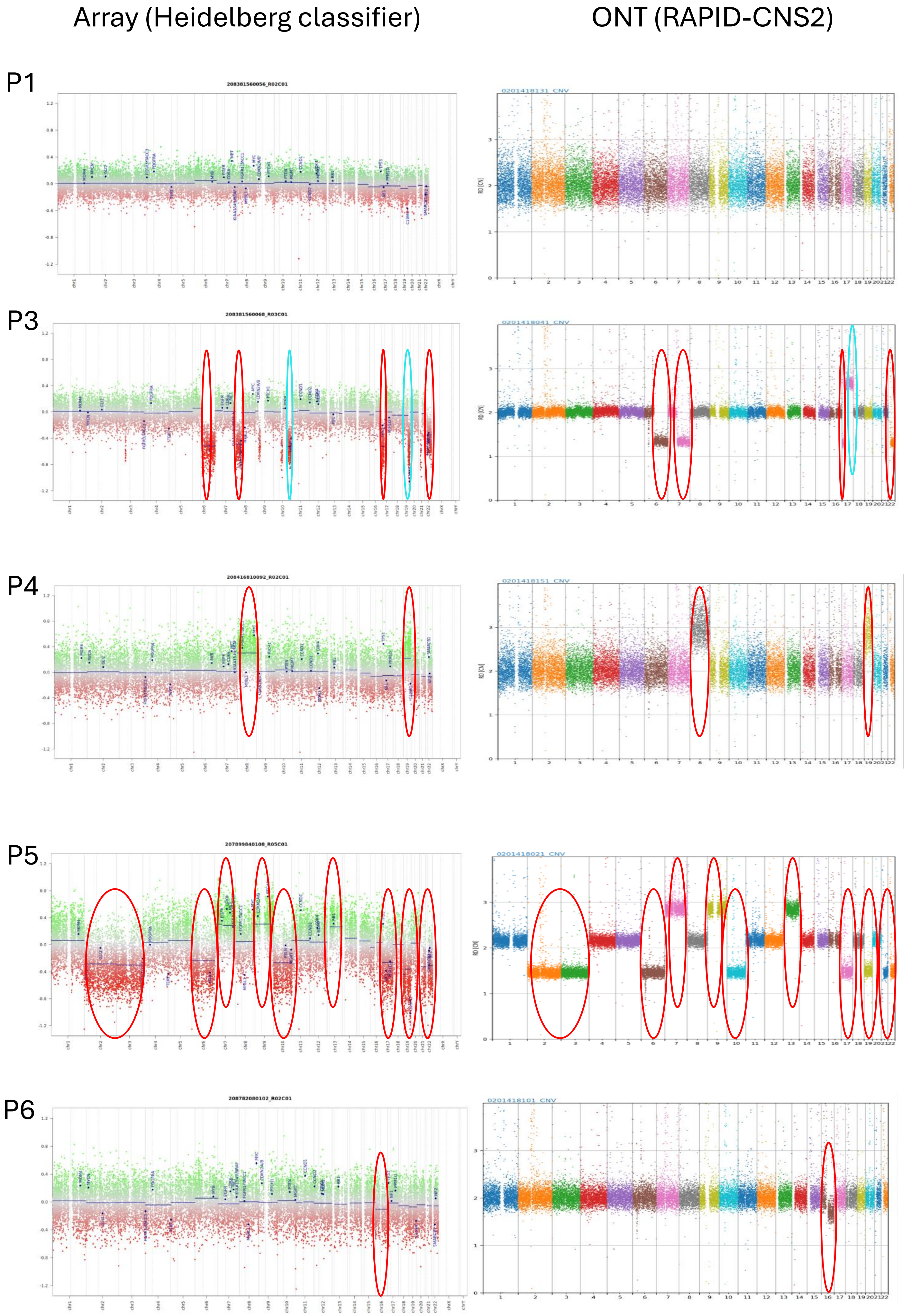

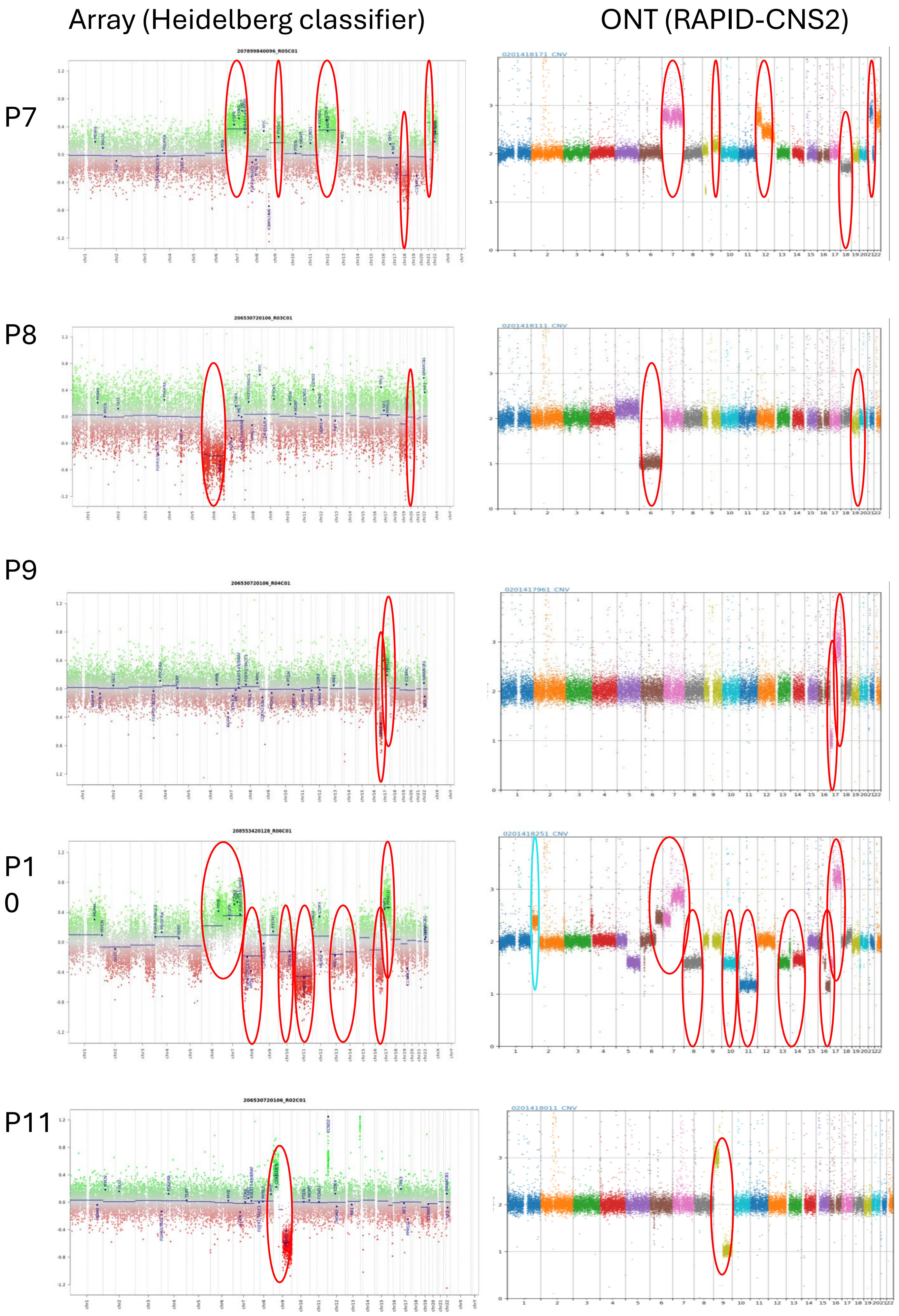

Array (Heidelberg classifier)
ONT (RAPID-CNS2)
P7
207899840096_R05C01
0201418171_CNV
P8
206530720106_R03C01
0201418111_CNV
P9
206530720106_R04C01
0201417961_CNV
P10
208553420128_R06C01
0201418251_CNV
P11
206530720106_R02C01
0201418011_CNV

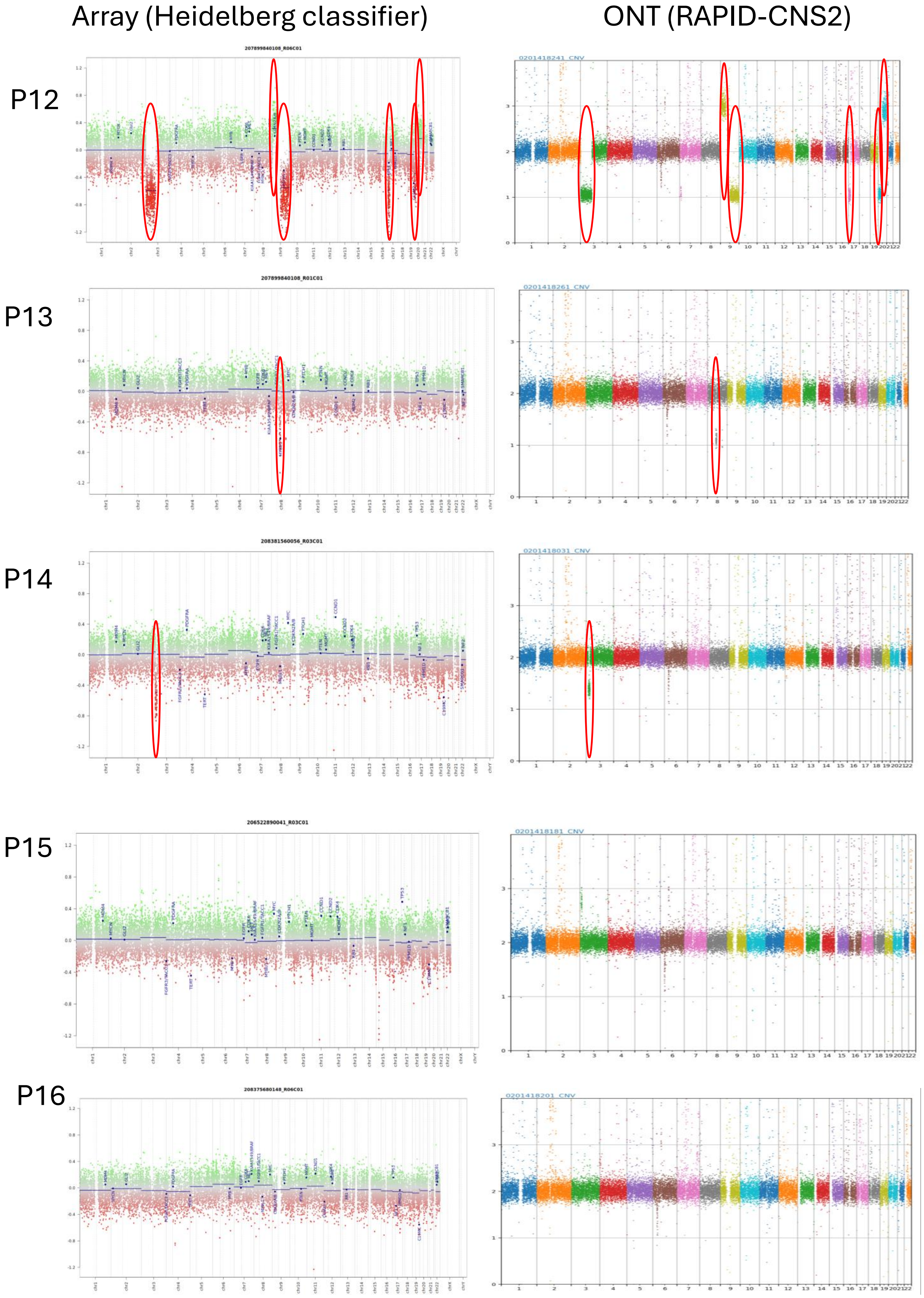
Array (Heidelberg classifier)
ONT (RAPID-CNS2)
P12
207899840108_R06C01
0201418241_CNV
P13
207899840108_R01C01
0201418261_CNV
P14
208381560056_R03C01
0201418031_CNV
P15
206522890041_R03C01
0201418181_CNV
P16
208375680148_R06C01
0201418201_CNV

Array (Heidelberg classifier) | ONT (RAPID-CNS2)

P17

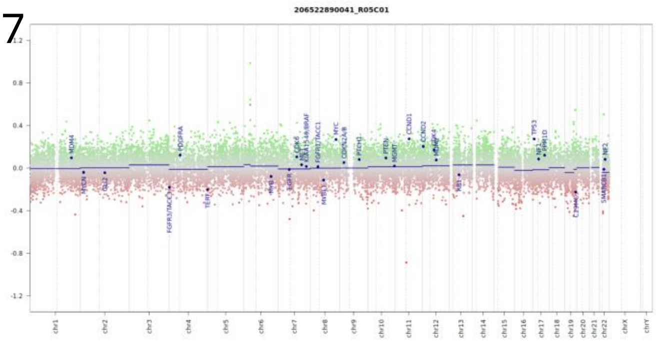


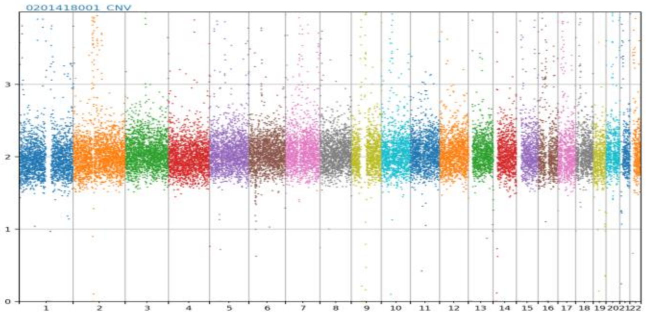


P18

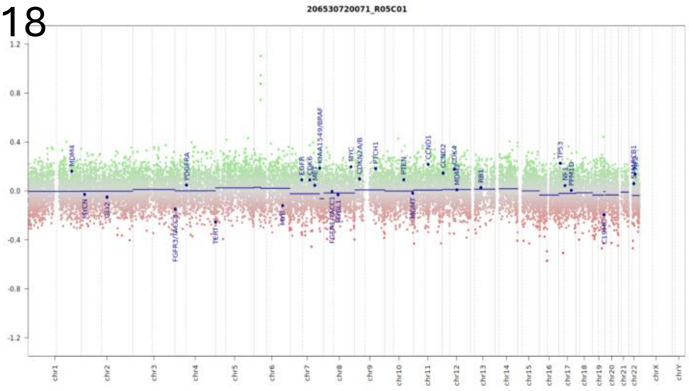


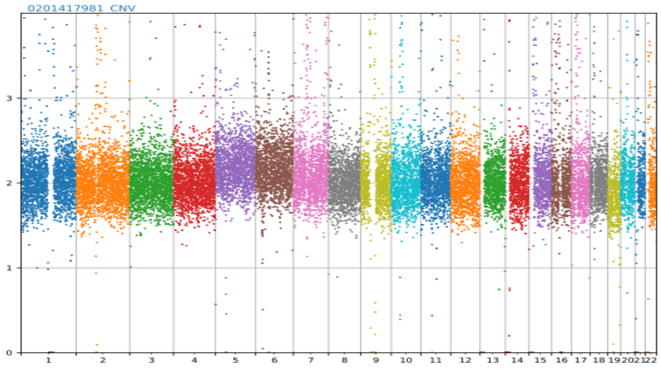

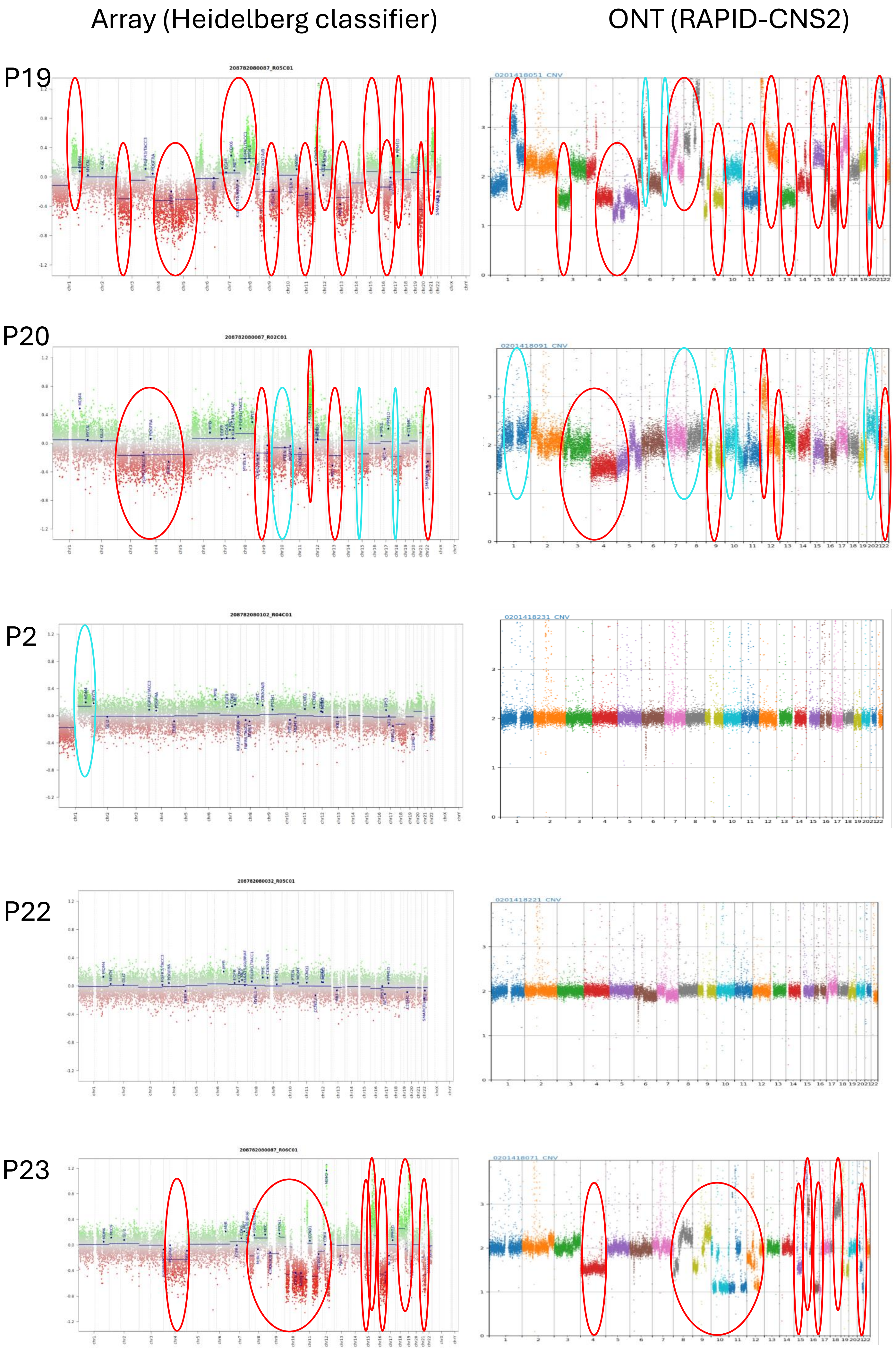
Array (Heidelberg classifier)
ONT (RAPID-CNS2)
P19
208782080087_R05C01
0201418051_CNV
P20
208782080087_R02C01
0201418091_CNV
P2
208782080102_R04C01
0201418231_CNV
P22
208782080032_R05C01
0201418221_CNV
P23
208782080087_R06C01
0201418071_CNV

Supplementary Figure 3

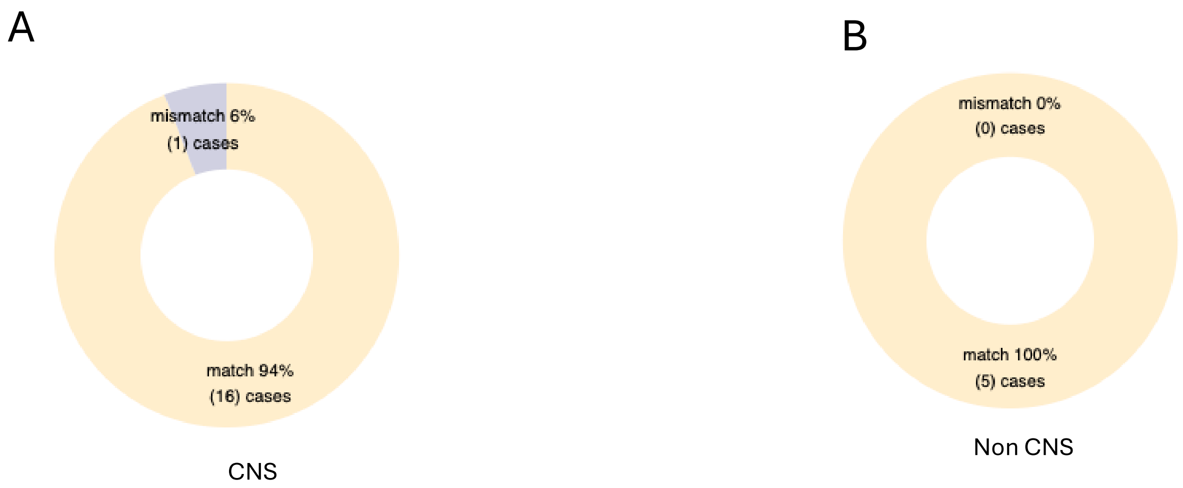
